# Reduced glymphatic function after traumatic brain injury measured using diffusion MRI

**DOI:** 10.1101/2022.11.05.22281969

**Authors:** Tracy Butler, Liangdong Zhou, Ilker Ozhasin, Xiuyuan Hugh Wang, Jacob Garretti, Henrik Zetterberg, Kai Blennow, Keith Jamison, Mony J. de Leon, Yi Li, Amy Kuceyeski, Sudhin A. Shah

## Abstract

The glymphatic system is a perivascular fluid clearance system, most active during sleep, considered important for clearing the brain of waste products and toxins. Glymphatic failure is hypothesized to underlie brain protein deposition in neurodegenerative disorders like Alzheimer’s Disease. Preclinical evidence suggests that a functioning glymphatic system is also essential for recovery from traumatic brain injury (TBI), which involves release of debris and toxic proteins that need to be cleared from the brain.

We estimated glymphatic clearance using Diffusion Tensor Imaging Along Perivascular Spaces (DTI-ALPS), an MRI-derived measure of water diffusivity surrounding veins, in 13 non-injured controls and 37 subjects with TBI (∼5 months post). We additionally measured plasma concentrations of Neurofilament light chain (NfL), a biomarker of injury severity, in a subset of subjects.

DTI-ALPS was significantly lower in TBI subjects compared to controls, after controlling for age, and significantly, negatively correlated with NfL.

Glymphatic impairment after TBI could be due to mechanisms such as mis-localization of glymphatic water channels, inflammation, proteinopathy and/or sleep disruption. Additional work, including longitudinal studies, are needed to confirm results and assess glymphatic associations with outcome. Understanding post-TBI glymphatic functioning could inform novel therapies to improve short-term recovery and reduce later risk of neurodegeneration.

## Introduction

Brain clearance can be broadly defined as the removal of waste from the brain via multiple, overlapping systems including active and passive transport at brain barriers, diffusion, and the glymphatic system.^1-4^ The glymphatic system involves subarachnoid cerebrospinal fluid (CSF) flowing into the brain alongside arteries, mixing with brain interstitial fluid (ISF) containing waste products, then flowing out of the brain along veins. This convective perivascular flow allows more rapid waste clearance than would be possible with diffusion alone. Impaired clearance has been hypothesized to be part of the pathophysiology underlying the buildup of the toxic proteins amyloid-β (Aβ) and tau causing Alzheimer’s disease (AD).^1, 4^

Clearance may also be important after traumatic brain injury (TBI), when there is neuronal debris and release of proteins including Aβ and tau which need to be cleared. TBI-induced clearance deficits may be one explanation for TBI increasing later risk for AD and other dementias.^5, 6^ In an animal model, glymphatic clearance disruption promoted tau deposition and impaired cognitive recovery.^7^ Understanding the role of clearance after TBI could provide novel therapeutic targets to enhance TBI recovery and reduce future risk of neurodegeneration.

Three prior studies have shown enlarged perivascular spaces (PVS) – the location where CSF mixes with ISF to facilitate clearance of waste – in TBI.^8-10^ This is considered evidence of possible glymphatic stasis and failed clearance.^11^ However, a static measure of PVS does not reflect the dynamic process of fluid flow. A new technique called diffusion tensor imaging along perivascular spaces (DTI-ALPS) quantifies actual water diffusion with PVS.^12, 13^ Specifically, DTI-ALPS allows evaluation of diffusivity parallel to PVS surrounding medullary veins at the level of the lateral ventricles. As shown in **Figure 1**, because the direction of PVS is orthogonal to projection and association white matter fiber tracks, axial diffusion at high b-value (to suppress intravascular venous flow) is uniquely sensitive to perivenous flow. ^12, 13^ Decreased glymphatic function was very recently demonstrated using this method in subjects with AD^14^ and TBI.^15^ Here we apply DTI-ALPS to assessing glymphatic function in subacute TBI and in relation to blood levels of neurofilament light protein (NFL) – a robust biomarker of TBI severity.^16, 17^

**Figure 1.**
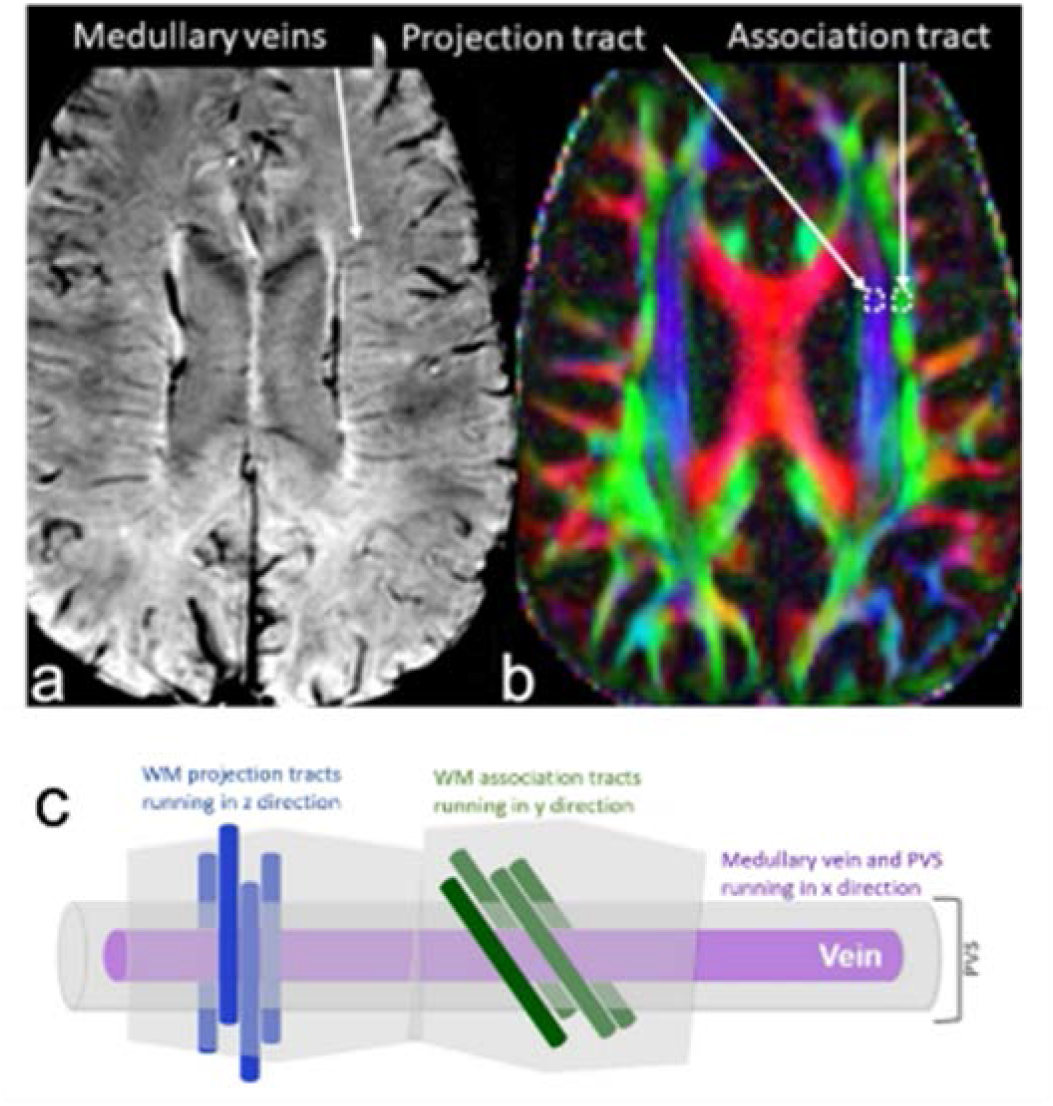
DTI-ALPS: Diffusion Tensor Imaging Along Perivascular Spaces. **(A)** Susceptibility weighted image showing clear medullary veins. **(B)** ROI placement within fiber tracts on color coded FA maps at the medullary veins. **(C)** A schematic adapted from Taoka et al.^13^ of the perivascular space running parallel to the medullary vein and therefore orthogonal to the principal fiber tract orientations.

## Materials and methods

TBI participants were recruited through inpatient rehabilitation units and trauma departments. Control subjects were recruited through local advertisements. All subjects provided informed consent prior to participation, and all study activities were approved by Weill Cornell Medicine’s Institutional Review Board. TBI subjects had sustained a complicated mild (Glasgow Coma Scale [GCS] score of 13–15 with evidence of intracranial lesion on acute neuroimaging) or moderate-severe TBI (GCS ≤ 12) within the prior 6 months. They were scanned ∼ 5 months after injury. Controls were free from medical and psychiatric illness and substance abuse.

### MRI data acquisition and processing

Anatomical T1w MPRAGE (0.8 mm isotropic) and multi-shell diffusion-weighted MRI (1.5 mm isotropic, b=1500,3000, 98 directions per shell) was acquired with a multi-band acceleration factor 4 on a 3T Siemens Prisma scanner with a 32-channel head coil. Diffusion data were collected with both anterior-posterior and posterior-anterior phase-encoding, with TE/TR=89.2/3230ms. DMRI-weighted data were corrected for susceptibility-induced geometric and eddy current distortions, and intervolume subject motion using the topup and eddy toolboxes.^18^ The preprocessed dMRI data were used to fit diffusion tensors and obtain fractional anisotropy (FA) and diffusivity maps for each subject in the directions of the *x*- (right-left,*D*_*xx*_), *y*- (anterior-posterior,*D*_*yy*_), and *z*-axes (inferior-superior,*D*_*zz*_).

*D*_*xx*_ corresponds to the direction of vessels in the periventricular white matter. Considering that the perivascular glymphatic system runs along these vessels, *D*_*xx*_ is assumed to reflect water diffusivity along the glymphatic system. As shown in **Figure 1**, each participant’s FA map was color coded in RGB style using the first eigenvector and 5-mm-diameter square ROIs were manually placed bilaterally in the projection area (predominantly in *z*-axis direction) and association areas (predominantly in *y*-axis direction) at the level of the lateral ventricle. The diffusivity values along the *x*-, *y*-, and *z*-axes within the ROIs were obtained for each participant. The ALPS index was calculated as a ratio of the mean of the *x*-axis diffusivity in the projection area (*D*_*xx, proj*_) and *x*-axis diffusivity in the association area (*D*_*xx, assoc*_) to the mean of the *y*-axis diffusivity in the projection area (*D*_*yy, proj*_) and the *z*-axis diffusivity in the association area (*D*_*zz, assoc*_).^12, 13^ No laterality difference were observed so left and right ALPS indices were averaged for data analysis. A higher ALPS index indicates greater diffusivity along PVS while an index close to 1.0 reflects minimal diffusion.

### NfL measurement

Plasma NfL concentration in blood drawn at the time of MR scanning was measured using single-molecule array (Simoa; Quanterix; Billerica, MA) performed at the Clinical Neurochemistry Laboratory, University of Gothenburg, Sweden in a single analytical run, following established methods.^16^

### Statistical Analyses

Analyses were performed in R (R Version 3.6.3). T-test and chi-square were used to assess group demographic and NfL differences. ANCOVA was used to compare the ALPS index between TBI subjects and controls while accounting for subject age. The relationship between ALPS index and plasma NfL was assessed using the following multivariate regression model: ALPS = *β*_0_ + *β*_1_ * NFL + *β*_2_ * Age.

## Results

### Demographics

Subject information is shown in **Table 1**. TBI subjects were non-significantly younger than controls (p=0.098). Males were over-represented in the TBI group as compared to controls, though this was not statistically significant (chi square = 2.183, p=0.14.)

**Table 1.** Participant demographics.

| **Table 1. Participant demographics.** |  |  |
| --- | --- | --- |
|  | **Subjects with TBI** | **Controls** |
| Sex | 28 male (75.7%); n=37 | 7 male (53.8%); n=13 |
| Age (mean, range)_ | 51.5 (range: 19-85, std=16.7); n=37 | 58.5 (range: 33-86, std=15.1); n=13 |
| Time since injury (mean, range) | 148.7 (range: 62-241, std=39); n=37 | N/A |
| Glasgow Coma Score (mean, range) | 12.3 (range: 5-15, std=2.9); n=17 | N/A |

### ALPS

As shown in **Figure 2**, ALPS index was significantly lower in subjects with TBI compared to controls after controlling for age: TBI mean = 1.336, std 0.155; control = 1.389, std .164; F[1,47]=4.083, p=0.049.

**Figure 2.**
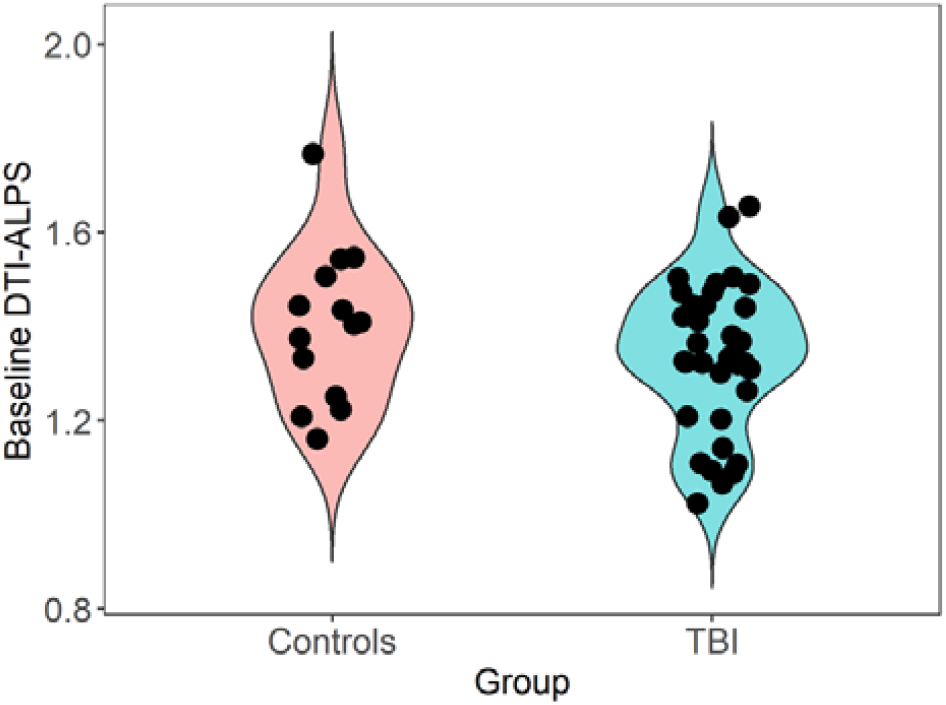
Violin plot showing distribution of ALPS-index in TBI subjects versus controls. When controlling for age, TBI subjects had significantly (p=0.049) lower ALPS index as compared to controls, consistent with glymphatic dysfunction.

### NfL

Subjects with TBI (n=17)_ had higher plasma NfL concentration than controls (n=9): TBI mean=90.59, std=115.18; control mean= 22.65, std=20.99; t = -2.72, p=0.012. As shown in **Figure 3**, ALPS index correlated significantly with NfL (beta = -0.725, p = 0.029) in subjects with TBI, wherein lower ALPS index was related to higher NfL. There was also a significant, negative correlation between ALPS and age (beta = -0.781, p = 0.020.)

**Figure 3.**
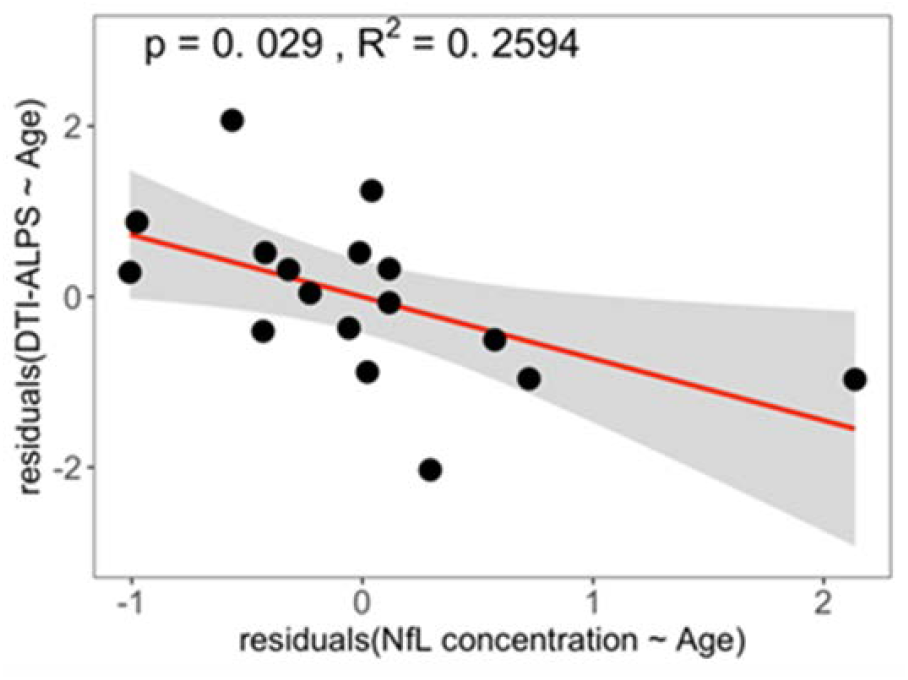
Partial regression plot of ALPS index with plasma NfL in 17 TBI subjects. Regression model: ALPS = β_0_ + β_1_ * NFL+ β_2_ * Age. Higher NfL, indicative of greater neural injury, is associated with lower glymphatic function.

## Discussion

ALPS index,^12, 13^ an indicator of the activity of the glymphatic system, was significantly lower in subacute TBI subjects compared to controls, even after controlling for effect of age. Results are in accord with one animal study^7^ and one recent clinical study.^15^ The correlation of ALPS index with plasma NfL – a biomarker of brain injury severity^16, 17^ – indicates that greater neuronal damage is associated with worse glymphatic function.

Mechanisms for decreased clearance after TBI may include sleep disruption, mis- localization of aquaporin-4 water channels involved in glymphatic clearance, protein deposition and sleep disruption.^7^ Impaired clearance after TBI - when cellular debris, blood, inflammatory cells and proteins including the pathognomic AD proteins AB and tau need to be cleared – would be expected to affect short term recovery after injury and contribute to later risk of proteinopathy and neurodegeneration.^19^ Additional study of post-TBI clearance in humans and animal models is warranted to clarify relevant mechanisms and guide targeted therapy. Because sleep is required for optimal clearance,^3^ and TBI is well known to impair sleep,^20^ focusing on improving sleep after TBI may be a promising strategy.^9^

ALPS index was affected by aging as well as TBI, with lower ALPS index in older subjects. Clearance is known to decline with aging.^1, 4^ Demonstration of significant group differences and correlation with NfL, after controlling for age, support DTI-ALPS as a sensitive marker of TBI-induced glymphatic deficit. Our results are consistent with recent work by Park et. al.^15^ in a similar population of subjects scanned ∼one month after TBI.

We interpret the negative correlation between ALPS and NfL as reflective of TBI severity, with lower ALPS and higher NfL both indicating greater neural damage. However, these two measures may be related. The movement of brain proteins used as TBI biomarkers from brain to blood depends upon clearance mechanisms including glymphatic function and blood-brain barrier breech to a variable degree. This introduces a potential confound, with blood biomarker levels reflecting both degree of injury and movement from brain to blood, which can be altered by that injury. In an animal model of TBI, impaired clearance has been shown to decrease blood levels of the TBI biomarkers S100β, GFAP and neuron specific enolase to control levels.^21^ However, NfL levels are not considered to be affected by alterations in the blood brain barrier,^22^ and current results suggest that if glymphatic dysfunction causes decreased blood levels of NFL, the effect may be minor. Additional studies including larger numbers of subjects and additional blood and neuroimaging biomarkers reflecting different aspects of clearance are greatly needed. Not accounting for clearance has been proposed as the answer to the question, “Why have we not yet developed a simple blood test for TBI?”^21^

This study has several limitations. We use DTI-ALPS to measure perivascular glymphatic clearance. However, there is significant controversy about details of fluid clearance, and it remains challenging to assess in both animal and humans.^1, 23^ Other methods for interrogating fluid clearance include PET measurements of radiotracer efflux from ventricles,^24, 25, 26^ fMRI measurement of coordinated neural and fluid pulsations linked to and driving CSF pulsations^27, 28^ and dynamic contrast MRI.^29^ Determining which method or combination of methods best reflects clearance in humans is hampered by the absence of a “gold standard” clearance measure. However, DTI-ALPS has significant face validity as a measure of *actual* diffusivity of fluid along PVS and has been shown to correlate with the rate at which intrathecally-injected contrast appears in perivascular spaces,^30^ considered one of the most direct measures of glymphatic clearance obtainable in humans.^31^ The TBI group was over 75% male, reflecting the epidemiology of moderate TBI.^32^ Larger studies including greater numbers of women are needed to assess possible sex differences in glymphatic function, which may be relevant to important sex differences in TBI mechanisms and outcomes.^33^

The ALPS index difference between TBI subjects and controls was modest; we hypothesize this may relate to the subacute nature of the MRI scan (∼5 months after injury). It is likely that scanning subjects in the acute phase of recovery will capture greater glymphatic impairment that resolves over time. Longitudinal analysis of clearance function changes after TBI are needed.

## Data Availability

All data produced in the present study are available upon reasonable request to the authors

